# Deep Learning-based Workflow for Automatic Extraction of Atria and Epicardial Adipose Tissue on cardiac Computed Tomography in Atrial Fibrillation

**DOI:** 10.1101/2023.05.03.23289448

**Authors:** Guan-Jie Wang, Ling Kuo, Shih-Lin Chang, Yenn-Jiang Lin, Fa-Po Chung, Li-Wei Lo, Yu-Feng Hu, Tze-Fan Chao, Ta-Chuan Tuan, Jo-Nan Liao, Ting-Yung Chang, Chin-Yu Lin, Chih-Min Liu, Shin-Huei Liu, Ming-Ren Kuo, Guan-Yi Lee, Yu-Shan Huang, Cheng-I Wu, Shih-Ann Chen, Chia-Feng Lu

## Abstract

**Background:** Preoperative measurements of left atrium (LA) and epicardial adipose tissue (EAT) volumes in computed tomography (CT) images have been reported to be associated with an increased risk of atrial fibrillation (AF) recurrence. We aimed to design a deep learning-based workflow to provide a reliable automatic segmentation of atria, pericardium and EAT, which can facilitate future applications in AF.

**Methods:** A total of 157 patients with AF who underwent radiofrequency catheter ablation were enrolled in this study. The 3D U-Net models of LA, right atrium (RA) and pericardium were used to develop the pipeline of total, LA-and RA-EAT automatic segmentation. We defined the attenuation range between -190 to -30 HU as fat within the pericardium to obtain total EAT. Regions between the dilated endocardial boundaries and endocardial walls of LA or RA within the pericardium were used to detect the voxels attributed to fat, resulting in LA-EAT and RA-EAT.

**Results:** The LA, RA and pericardium segmentation models achieved Dice coefficients of 0.960 ± 0.010, 0.945 ± 0.013 and 0.967 ± 0.006, respectively. The 3D segmentation models correlated well with ground truth for LA, RA and pericardium (r=0.99 and p < 0.001 for all). For the results of EAT, LA-EAT and RA-EAT segmentation, Dice coefficients of our proposed method were 0.870 ± 0.027, 0.846 ± 0.057 and 0.841 ± 0.071, respectively.

**Conclusions:** Our proposed workflow for automatic LA/RA and EAT segmentation applying 3D U-Nets on CT images was reliable in patients with AF.

## Introduction

Atrial fibrillation (AF) is the most common cardiac arrhythmia, which may alter the morphology and function of the left atrium (LA) and the epicardial adipose tissue (EAT) [1, 2]. Prior to the treatment of AF, pulmonary vein computed tomography (CT), which is a type of contrast-enhanced cardiac CT, is commonly used to provide safe and successful ablation procedures [3]. Cardiac CT imaging is the gold standard technique to measure the atrial and EAT volumes because it can achieve high spatial resolution and whole-heart coverage [4]. Many studies have shown that LA [5-7], right atrial (RA) [8, 9], total EAT [10, 11] and LA-EAT volumes [11, 12] measured in CT images are important features for the prediction of AF recurrence after catheter ablation.

In clinical practice, cardiologists often apply the manual delineation for atrial and EAT segmentation to obtain volume measurements. However, the manual segmentation is extremely time-consuming and subjective. The automatic segmentation of atria and EAT on cardiac CT images using deep learning is valuable in improving consistency and efficiency. Many studies have designed different network architectures to extract whole heart structures, including LA, RA, left ventricle (LV), right ventricle (RV) and LV myocardium on cardiac CT dataset [13-16]. However, in these studies for whole heart segmentation, the datasets used to develop the deep learning models were not collected for a specific disease such as AF. Additionally, because AF may lead to structural remodeling, including LA dilatation and tissue fibrosis [1], reliable automatic segmentation of cardiac structures may not be achieved in previously proposed deep learning models. The recent studies have developed deep learning models for LA automatic segmentation in AF patients [17, 18]. However, in patients with AF, we have to perform a comprehensive segmentation system, including atria, pericardium and EAT, to predict AF recurrence after catheter ablation. There are no current studies proposing a complete workflow for automatic segmentation of cardiac structures associated with AF recurrence to provide a convenient system for clinical use. For total EAT segmentation, although some semi-automatic methods were proposed to identify Hounsfield unit (HU) corresponding to fat, these methods still required to manually delineate the pericardium as the outer boundary, which could not prevent the time spent on contouring and inter-observer variability [19, 20]. Furthermore, recent studies have proposed the extraction of total EAT using deep learning algorithms in non-contrast or contrast-enhanced cardiac CT images, but no studies have developed the automatic segmentation of LA- and RA-EAT [11, 12, 21-23].

In this study, we aimed to provide a new workflow to automatically segment atria, pericardium and EAT on contrast-enhanced cardiac CT images in AF patients. The automatic segmentation in this system based on deep learning could achieve reliable results, improve the work efficiency, and contribute to the future image studies in AF.

## Materials and Methods

### 2.1. Dataset for development of auto-segmentation models

We retrospectively collected the contrast-enhanced cardiac CT images of 157 patients with AF from Taipei Veterans General Hospital (TVGH). The cardiac CT scan was used to assess the morphology of LA and pulmonary veins prior to the procedures for radiofrequency catheter ablation. Inclusion criteria were: (1) sufficient image quality for evaluation of LA and pulmonary veins; (2) no metal implants in the cardiac chambers; and (3) presence of whole heart within the image volume. This study was approved by the Institutional Review Board of TVGH (VGH-IRB Number: 2022-06-016AC), and the requirement of informed consent was waived.

### 2.2. Cardiac CT acquisition

Cardiac CT scans from TVGH were acquired on 64-slice (Aquilion 64, Toshiba Medical Systems) and 256-slice (Brilliance iCT, Philips Healthcare) scanners. Aquilion 64 used the peak tube voltage of 100 kVp and tube current of 350 mA, whereas Brilliance iCT used the peak tube voltage of 100 or 120 kVp and tube current of 596 mA. The 16-bit and 12-bit grayscale PVCT images were generated by Aquilion 64 and Brilliance iCT, respectively. Cardiac CT images from each patient were reconstructed using a slice thickness of 1 mm and stored in DICOM format with a matrix size of 512 × 512. The variation of pixel sizes for reconstructed images was 0.34 × 0.34 to 0.72 × 0.72 mm^2^.

### 2.3. Manual labeling

The LA, RA and pericardium were manually contoured using the Multimodal Radiomics Platform (http://cflu.lab.nycu.edu.tw/MRP_MLinglioma.html), which was developed in the MATLAB environment [24]. Cardiac CT images viewed on this platform were displayed with a soft tissue window (window center: 40 HU, window width: 400 HU) to improve the contrast of all cardiac structures. The region of interest (ROIs) for LA and RA, including the atrial appendages, were contoured by tracing the atrial endocardial boundaries. The upper border of the pericardium was defined as the top slice of the LA, while the inferior border was defined at the last slice containing any part of the cardiac chambers [25]. All ROIs were delineated by an experienced radiologic technologist and a cardiologist.

### 2.4. Image preprocessing

The full-range cardiac CT and ROI images were performed image pre-processing. First, the adjustments of the image resolution were resampled into the isotropic voxels of 0.5 × 0.5 × 0.5 mm^3^. Second, the z direction of all images was adjusted to the same slice size. In the cardiac CT images, we found the aortic arch as the initial slice and included the downward 320 slices to encompass the whole heart. If the patients had a large cardiac morphology, the initial slice would move down until the whole heart was covered. Third, the images were cropped to 400 × 400 and ensured that the ROIs of LA, RA and pericardium were included in the matrix. The CT images were further normalized to the soft tissue window and subsequently rescaled to an 8-bit grayscale from 0-255. Finally, the matrix size of the CT and ROI images was 400 × 400 × 320, with the isotropic voxels of 0.5 × 0.5 × 0.5 mm^3^. The input size of 3D U-Net was further downsampled to 200 × 200 × 160 with an interpolation of the voxel size to 1 × 1 × 1 mm^3^. All image pre-processing steps were performed in Matlab 2020a.

### 2.5. Deep learning models

To make the training set sufficient for three-dimensional (3D) segmentation, we had to prepare hundreds of labeled images. However, manual delineation of LA, RA and pericardium for all AF patients in the dataset was a laborious work. Hence, we used two-dimensional (2D) U-Nets trained by 30 patients including 9600 images to assist in manual labeling. The architecture and training parameters of 2D U-Nets were shown in **Supplementary materials**. The results of 2D U-Net segmentation for LA, RA and pericardium were modified by erasing or filling in the incorrect regions. In the 3D segmentation, the dataset consisted of 157 patients divided into 125 patients as the training set and 32 patients as the testing set. For the improvement of predictive ability, data augmentation was applied to increase the size and variability of training set [26]. The training images were rotated by 10, 20 and -20 degrees, resulting in 500 image volumes being used as the new training set.

Our proposed 3D U-Net architecture consisted of 22 convolutional layers with concatenation between the encoders and decoders. The encoders comprised repeated application of three convolutional layers with 3 × 3 × 3 kernels, each followed by batch normalization and ReLU processes. After every three convolutions, a 2 × 2 × 2 max-pooling with a stride of 2 was applied in succession. Each decoder consisted of a 2 × 2 × 2 transposed convolutional kernel with a stride of 2 and three 3 × 3 × 3 convolutional layers, each followed by batch normalization and ReLU. Finally, a 1 × 1 × 1 convolutional layer with softmax function was used to predict probability of each pixel. Our proposed 3D U-Net architecture was displayed in **Figure 1**.

**Figure 1.**
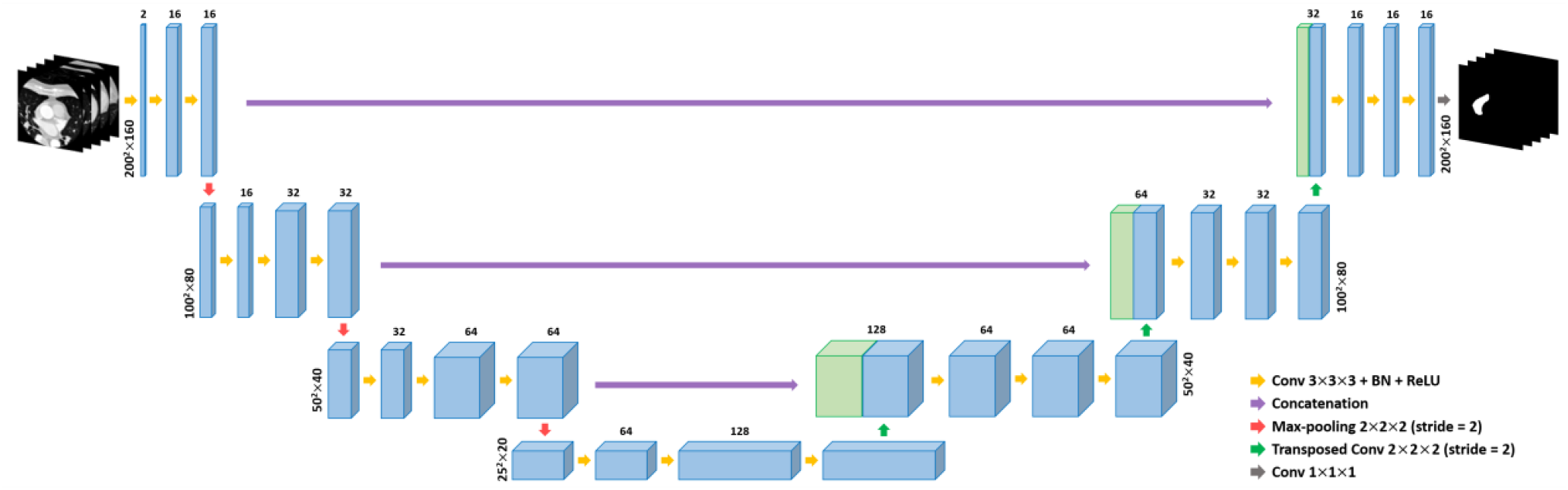
3D U-Net architecture. U-Net is the most famous fully convolutional neural network for satisfactory biomedical image segmentation. Three independent 3D U-Net models for LA, RA and pericardium segmentation were developed.

The experimental environment was implemented using MATLAB 2020a with a 6 GB NVIDIA GeForce RTX 2060 GPU, an Intel Core i7-9700 CPU and 40 GB of RAM. Training used the stochastic gradient descent with momentum of 0.9 to update the weight of the network. Training parameters for 3D U-Net models were: learning rate = 0.01, mini-batch size = 2, max epoch = 32, learning rate drop period = 10, and learning rate drop factor = 0.3. Dice loss was used as the loss function. It took approximately 52 hours and 50 minutes to complete the training of the models for LA, RA and pericardium segmentation, respectively.

### 2.6. EAT segmentation

EAT is the visceral fat deposited between the myocardium and the pericardium [27]. After automatically segmenting the pericardium by 3D U-Net (**Figure 2A**), cardiac CT images were resampled to the original voxel size of 0.5 × 0.5 × 0.5 mm^3^ and the voxels corresponding to EAT were identified. Because cardiac CT scan was performed with the contrast injection, a higher HU ranging from -190 to -30 was used for identifying the voxels within the pericardium [28, 29]. Sum of all these detected voxels within the pericardium were defined as the total EAT (**Figure 2B**).

**Figure 2.**
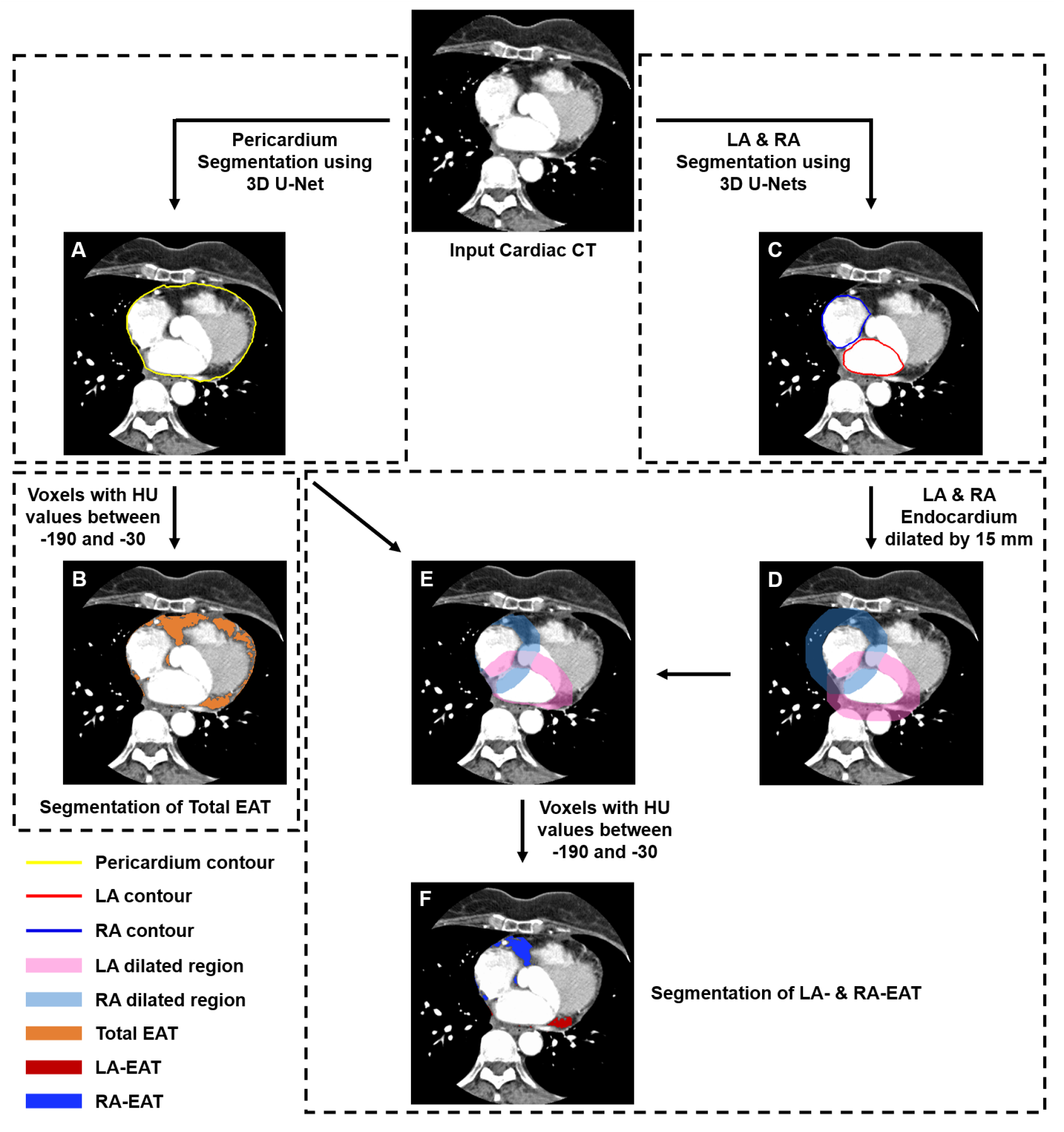
Flowchart of total EAT, LA-EAT and RA-EAT segmentation. Figure A shows the results of 3D pericardium segmentation. Figure B presents the results of total EAT segmentation within the pericardium, which identifies voxels with HU values between -190 and -30. Figure C shows the results of 3D LA and RA segmentation. Combining the steps of D, E and F yielded the results of LA- and RA-EAT segmentation.

The LA- and RA-EAT were the fat accumulated around LA and RA, respectively. LA and RA contours were extracted from the LA and RA ROIs segmented by our proposed 3D U-Nets (**Figure 2C**). According to a previous study, Wang et al. found that the mean thickness of the left and right atrioventricular groove EAT was 12.7 and 13.9 mm, respectively [30]. Because of this evidence, the endocardial boundaries of LA and RA were dilated by 15 mm individually to identify the fat surrounding both atria.

The areas for identifying the voxels of LA-EAT and RA-EAT were between the endocardial wall and the dilated boundaries (**Figure 2D**). We further automatically segmented the ROIs within the pericardium to exclude the extrapericardial fat (**Figure 2A**). Pixels between -190 and -30 HU could be found at the intersection of the dilated regions and the pericardium ROIs (**Figure 2E**). Because some pixels were part of both LA-EAT and RA-EAT, the reassignment of all pixels was performed. The minimum Euclidean distance was applied to measure the distance between each EAT pixel and both atria, which is defined as follows:

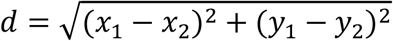

Where d is the minimum distance and the coordinates of EAT pixel and pixel situated at the atrial endocardial boundary are (*x*_1_,*y* _1_) and (*x*_2_,*y* _2_). When the minimum Euclidean distance of a pixel as EAT from LA was shorter than the distance from RA, the pixel would be considered as LA-EAT, and vice versa (**Figure 2F**). When the minimum distance of an EAT pixel from the LA was equal to the distance from the RA, this EAT pixel would be shared by LA- and RA-EAT.

### 2.7. Assessment of model performance

For the comparison of the similarity between manual and automatic segmentation, we used Dice coefficient, sensitivity and precision as statistical evaluation metrics [31], can be defined as follows:

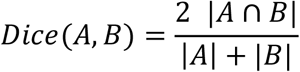

Where A is the automatic segmentation and B is the manual segmentation. Dice coefficient is between 0 and 1, with 0 indicating no overlap of segmentation and 1 indicating perfect overlap of segmentation.

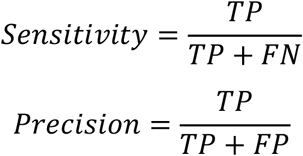

Where the overlapping pixels are classified into four categories, including true positive (TP), true negative (TN), false positive (FP) and false negative (FN).

We further measured percentage volume difference (PVD) to compare the differences between manual and automatic segmentation, which is based on following formula:

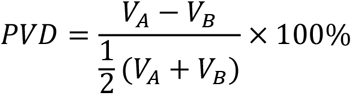

Where V_A_ is the volume of automatic segmentation and V_B_ is the volume of manual segmentation. The volume measurements were calculated by multiplying the voxel size by the sum of the voxels corresponding to each label.

### 2.8. Statistical analysis

Continuous variables including Dice coefficient, sensitivity, precision and volume difference were stated as mean ± standard deviation. The volume difference between automatic and manual segmentation was assessed by single-tailed paired t-tests. Bland-Altman plots and Pearson correlation coefficients were used to estimate for the bias and levels of agreement between automatic and manual segmentation. Statistical significance for all comparisons was accepted at p value < 0.05. Statistical analysis was conducted using Matlab 2020a.

## Results

### 3.1. Performance of auto-segmentation for LA, RA and pericardium

Comparison and evaluation metrics of ground truth and automatic segmentation with 2D U-Net assistance were shown in **Supplementary materials**. With the assistance of 2D U-Net, doctors could obtain initial 3D contours of LA, RA and pericardium to further apply manual revision, especially pulmonary vein-LA junctions, RA-superior vena cava junctions, RA-inferior vena cava junctions and pericardium of inferior ventricle.

The final 3D U-Net segmentation for LA, RA and pericardium took about 7.42 seconds per patient. For 3D U-Net models, the resultant LA, RA and pericardium segmentation achieved Dice coefficients of 0.960 ± 0.010, 0.945 ± 0.013 and 0.967 ± 0.006, respectively. The sensitivity obtained in 3D U-Net models of LA, RA and pericardium was 0.946 ± 0.023, 0.938 ± 0.026 and 0.965 ± 0.014, whereas the precision was 0.974 ± 0.013, 0.954 ± 0.024 and 0.969 ± 0.010, respectively. **Supplementary Figure S3** displayed the box plots of the Dice coefficients, sensitivity and precision for LA, RA and pericardium when using the testing dataset from 32 patients. **Figure 3** showed the visual assessment of 3D segmentation for LA, RA and pericardium.

**Figure 3.**
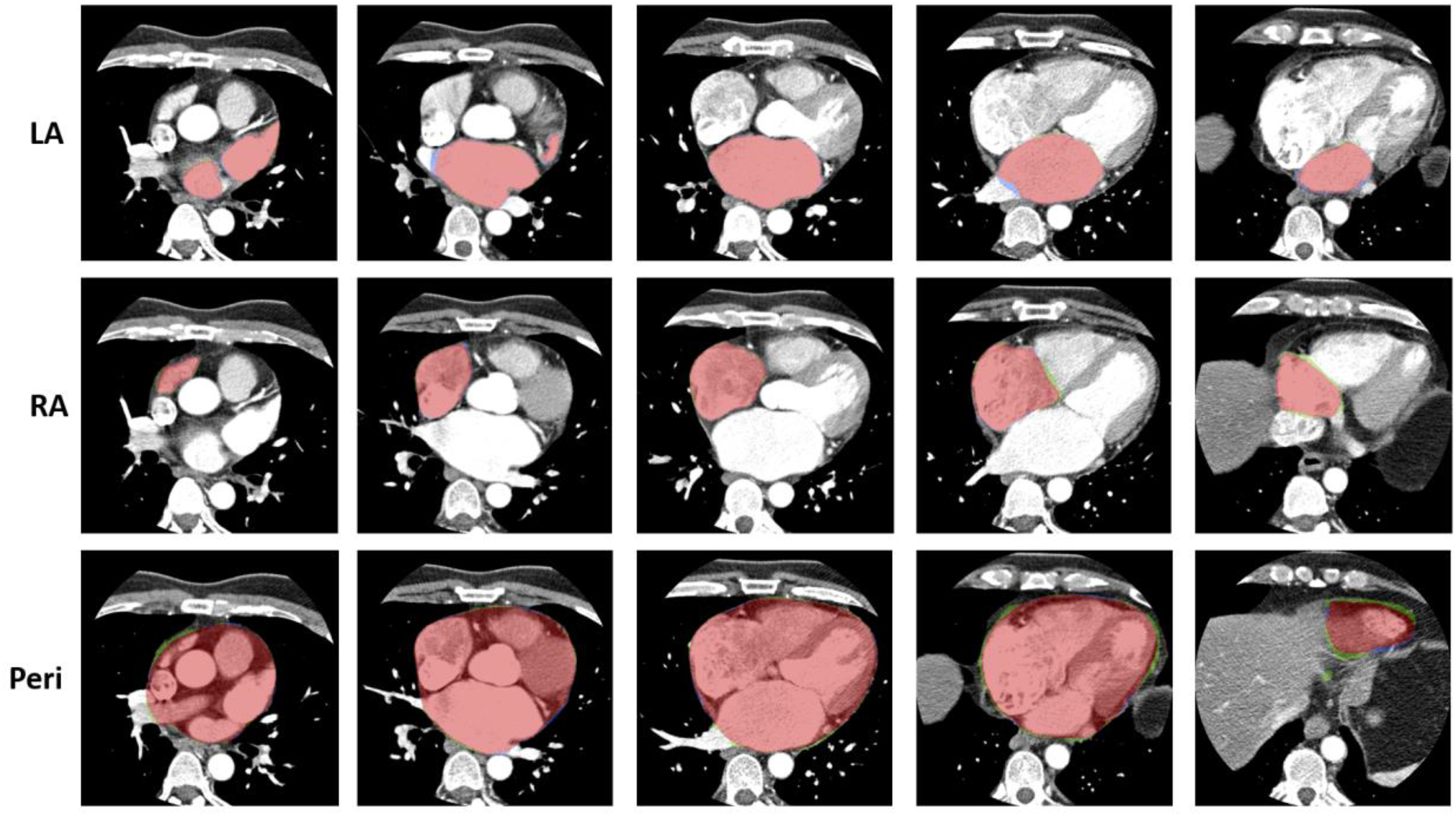
Ground truth and 3D U-Net segmentation of LA, RA and pericardium. The blue and green colors represent the ground truth and automatic segmentation of 3D U-Nets. The red color is the overlapping regions of ground truth and the automatic segmentation. Abbreviations: Peri = Pericardium.

The agreement and correlation of cardiac volumes obtained by 3D U-Nets and manual segmentation were shown in **Figure 4**. Comparing the volumes of LA, RA and pericardium extracted by manual and automatic segmentation, Bland-Altman plots presented the volume difference of -4.01 ± 4.62 mL, -2.34 ± 5.85 mL and -3.73 ± 18.80 mL, whereas the PVD of, -3.07 ± 3.35%, -1.89 ± 4.63% and -0.50 ± 2.24%, respectively. Volumes of LA (p<0.001) and RA (p=0.016) segmented by 3D U-Nets were significantly smaller than manual segmentation. For the segmentation of the pericardium, there was no significant volume difference between manual and automatic segmentation (p=0.135). By performing Pearson correlation analysis, the measurements of manual and automatic segmentation showed a significantly high correlation in all cardiac structures (r=0.99 for LA, RA and pericardium, p<0.001).

**Figure 4.**
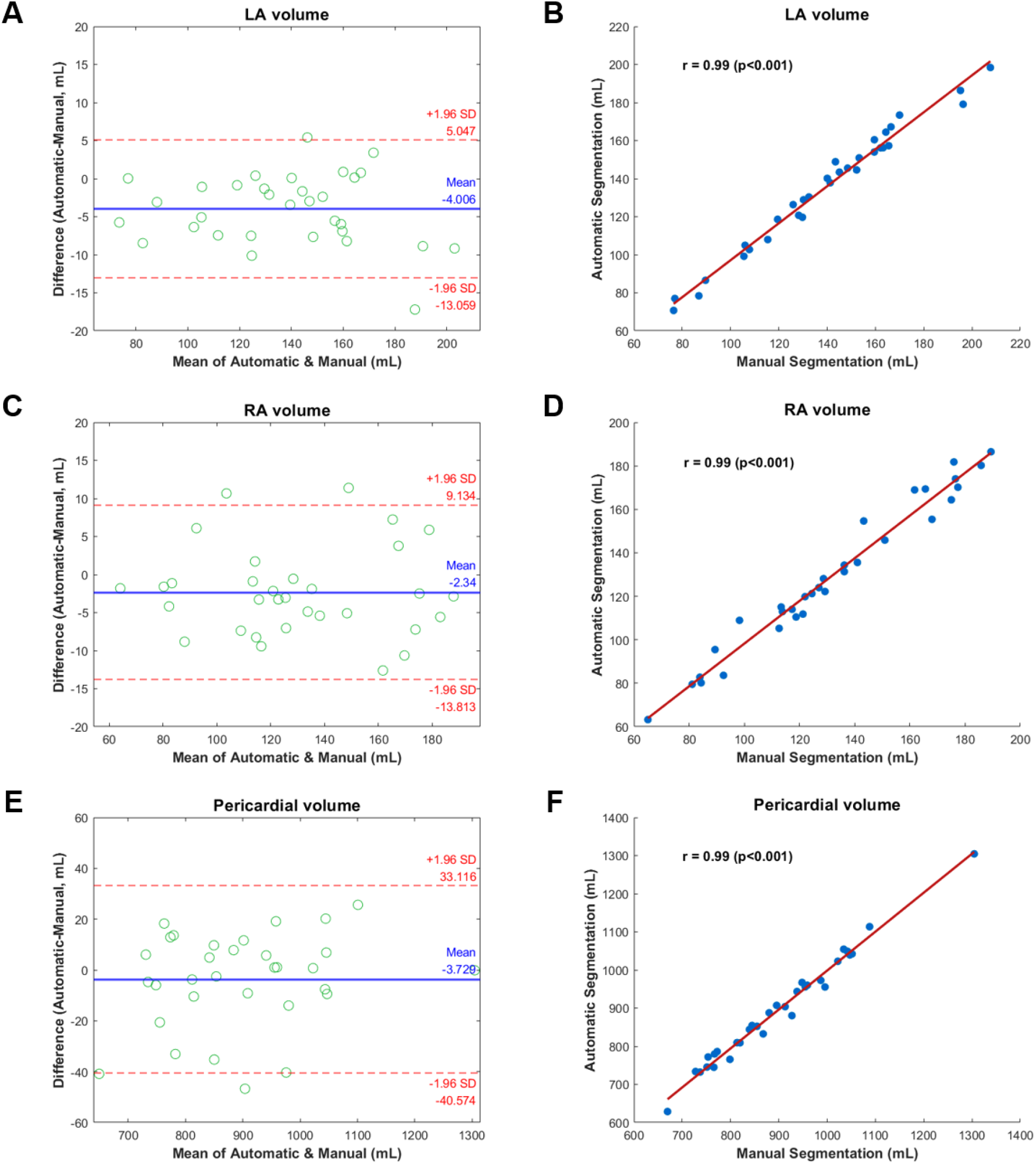
Comparison of the agreement for 3D automatic segmentation with manual segmentation of LA, RA and pericardium volumes. Bland-Altman analysis between our proposed 3D segmentation models and ground truth for LA (A), RA (C) and pericardium (E) volumes show mean bias (95% limits of agreement) of -4.01 (−13.06, 5.05) mL, -2.34 (−13.81, 9.13) mL and -3.73 (−40.57, 33.12) mL, respectively. Excellent correlations are found in 3D U-Net models against ground truth for LA (B), RA (D) and pericardium (F), all indicating r=0.99 (p < 0.001). In Bland-Altman plots, blue line indicates the mean difference and the two red dashed lines indicate the limits of agreement, from -1.96 to +1.96 standard deviations.

### 3.2. Performance of auto-segmentation for EAT segmentation

The performance of total EAT segmentation within pericardial regions showed the Dice coefficient of 0.870 ± 0.027, sensitivity of 0.888 ± 0.033 and precision of 0.856 ± 0.052, respectively. For the performance of LA- and RA-EAT segmentation, our proposed method yielded a Dice coefficient of 0.846 ± 0.057, a sensitivity of 0.861 ± 0.051 and a precision of 0.836 ± 0.082 for LA-EAT segmentation, and a Dice coefficient of 0.841 ± 0.071, a sensitivity of 0.839 ± 0.070 and a precision of 0.846 ± 0.085 for RA-EAT segmentation. The EAT segmentation including total EAT, LA-EAT and RA-EAT took about 31.35 seconds per patient. LA-EAT was mainly located in three regions: (a) areas surrounded by the ascending aorta, right superior pulmonary vein and superior vena cava; (b) those between the left atrial appendage and pulmonary trunk, and (c) those within the left atrioventricular groove. RA-EAT was primarily distributed in the right atrioventricular groove. A small part of RA-EAT was located in areas posterior to RA and between RA and coronary sinus. Results of automatic segmentation for total EAT, LA-EAT and RA-EAT were shown in **Figure 5**.

**Figure 5.**
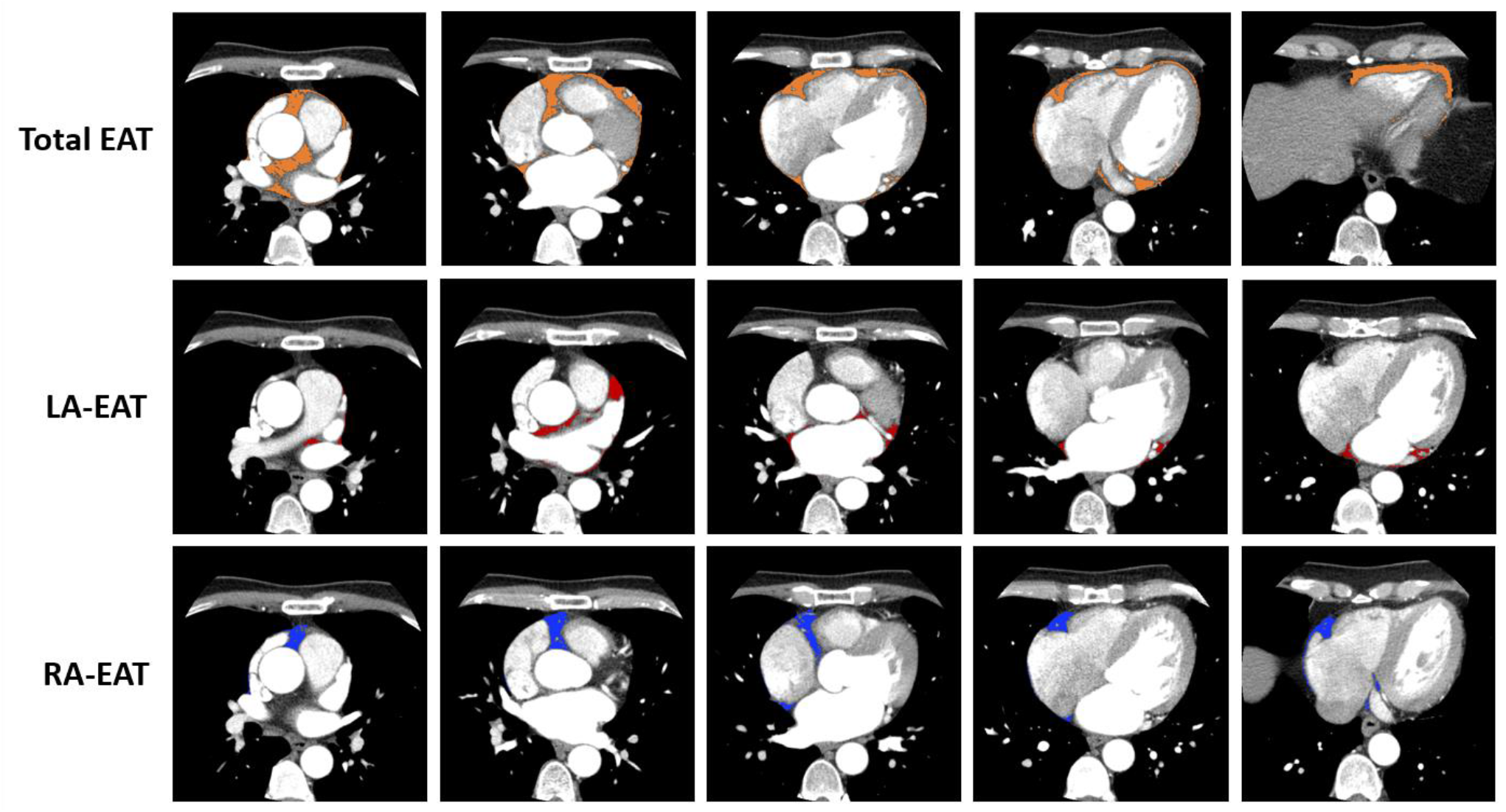
Distribution of total EAT, LA-EAT and RA-EAT in different CT axial slices. The brown, red and blue regions represent the total EAT, LA-EAT and RA-EAT identifying HU values between -190 and -30, respectively.

## Discussion

In this study, we proposed a full workflow based on deep learning to provide a reliable and rapid automatic segmentation for atria, pericardium and EAT on contrast-enhanced cardiac CT images in AF patients. In clinical practice, an experienced cardiologist would need to spend approximately 3 hours to manually delineate the atria and total EAT. In this study, we only required nearly 30 seconds to obtain the LA, RA, pericardium and EAT, consisting of total EAT, LA-EAT and RA-EAT, in patients with AF. Hence, this workflow can provide a time-efficient automatic segmentation for facilitating future applications in AF recurrence.

We summarized the methods and performance of our proposed models and following described articles for cardiac and EAT segmentation in **Table 1**. The LA volume measured in preoperative contrast-enhanced CT scan has been demonstrated to be an important feature in predicting AF recurrence [5-7], thus some studies developed the LA segmentation using deep learning on CT images in AF patients. Two studies employed a pipeline of two 2D convolutional neural networks (CNN) in AF patients, the first CNN for LA detection and the second CNN for LA segmentation [17, 18]. The study proposed by Chen et al. achieved an intersection over union (IoU) of 0.914 [17]; the study proposed by Abdulkareem et al. obtained a Dice coefficient of 0.885 ± 0.12 [18], respectively. Compared to the studies using 2D approaches described above, the present study achieved superior performance in LA segmentation (Dice coefficient = 0.960 ± 0.010) because 3D segmentation could provide global contextual information from volumetric CT images, particularly assisting in separating the boundaries of the LA and pulmonary veins. Additionally, our proposed workflow could provide the segmentation of other significant AF recurrence predictors, such as RA [8, 9], total EAT [10, 11] and LA-EAT [11, 12], to conduct a comprehensive assessment for AF patients undergoing catheter ablation.

**Table 1.**
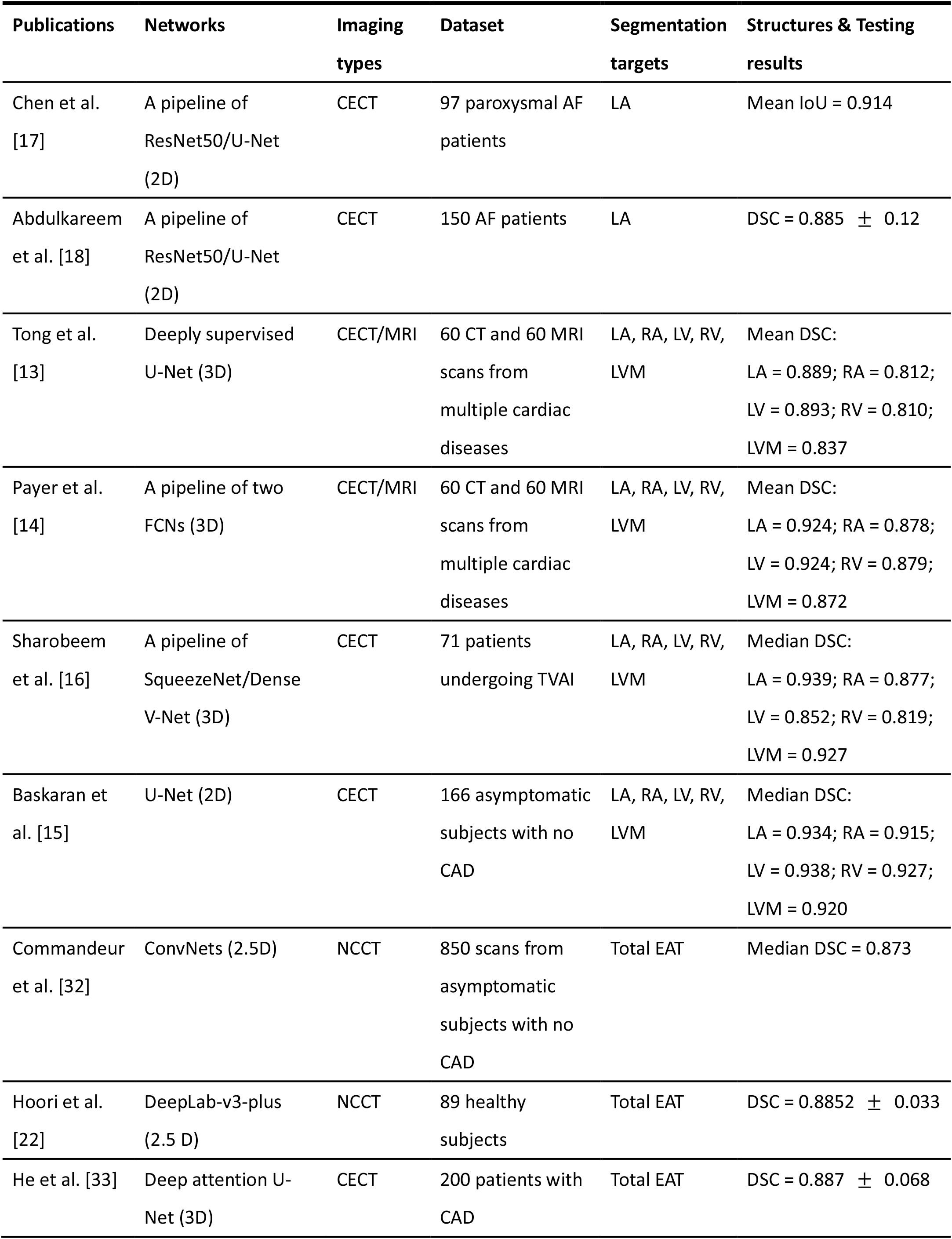
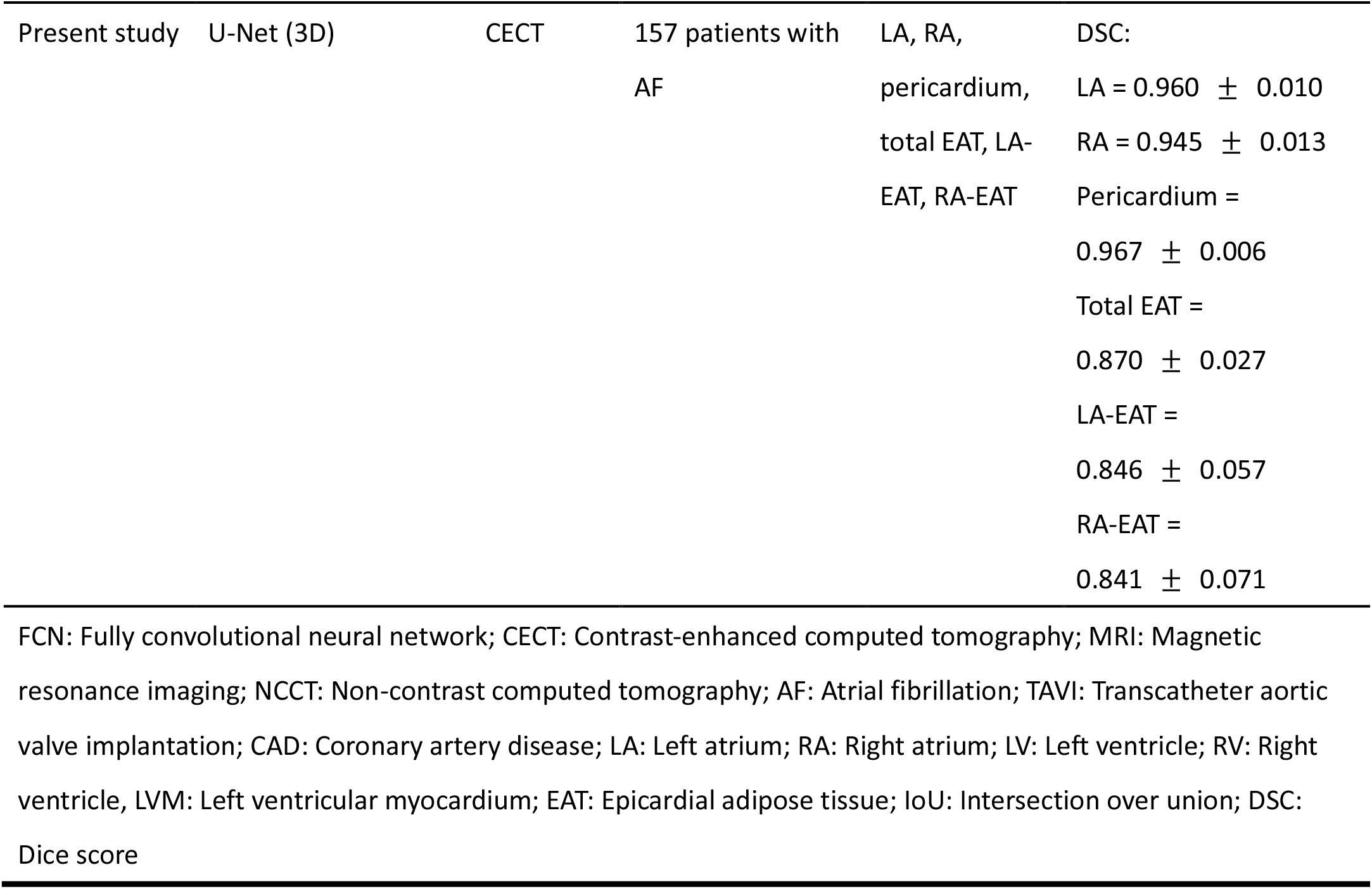
Related publications on cardiac CT and EAT segmentation using deep learning.

Several previous studies have presented the whole-heart segmentation using 3D deep learning methods, including LA, RA, LV, RV, LV myocardium, etc. The LA and RA segmentation in these studies achieved Dice coefficients ranging from 0.889-0.939 and 0.812-0.878, respectively [13, 14, 16]. Because our study used a dataset including more CT scans with sufficient variability in cardiac morphology, the superior performance in LA and RA segmentation (LA: 0.960 ± 0.010; RA: 0.945 ± 0.013) was obtained compared to these studies. To make segmentation models practical and meaningful, our study also evaluated the volume difference and correlation between manual and automatic segmentation. Although the LA and RA volumes of our proposed 3D segmentation were significantly smaller than the manual segmentation, the PVD of LA and RA was clinically acceptable in the range of 2-3%. A previous study proposed by Baskaran et al. used the 2D U-Net and obtained a great correlation in RA segmentation (r=0.97), but a slightly lower correlation in LA segmentation (r=0.78) [15]. That may because it was difficult to segment the blur junctions between the contrast-filled LA and LV by 2D segmentation. Compared to present study, our developed 3D models could provide completely contextual information and achieved the high correlation coefficients in LA (r=0.99) and RA (r=0.99) segmentation. Hence, we suggested that the 3D U-Net with an input of volumetric data outperformed the 2D U-Net with a single slice input for segmenting the complex structures of heart.

In the present study, we used contrast-enhanced cardiac CT images for performing pericardial and total EAT segmentation. The major advantage of contrast-enhanced CT is that it is easier to detect the pericardium compared to non-contrast CT. Although Commandeur et al. included 850 non-contrast CT scans from multiple cohorts to develop a fully-automatic approach for total EAT segmentation, they obtained a comparable result (median Dice=0.873) to the present study (median Dice=0.874) including 157 contrast-enhanced CT scans [32]. This proved that a large dataset had to be built to learn how to automatically segment the total EAT within the pericardium well in non-contrast CT images. Additionally, although Hoori et al. used the transfer learning with a bisect method to segment total EAT within the pericardium in non-contrast CT with a Dice coefficient of 0.8852 ± 0.033 [22], the total EAT segmentation in the present study using contrast-enhanced CT only required a simple 3D U-Net. In another previous study, the total EAT segmentation proposed by He et al. also used the contrast-enhanced CT to develop a 3D deep attention U-Net and obtained a similar mean Dice coefficient (0.887) to our study (0.870) [33]. However, the present study had a smaller standard deviation (0.027) of the Dice coefficient compared to the study proposed by He et al. (standard deviation with 0.068), which meant that our developed method was superior.

In this study, we first proposed the automatic segmentation of LA- and RA-EAT by combining 3D U-Nets of LA, RA, and pericardium. We noted that the Dice coefficients for LA- and RA-EAT were slightly lower than those for LA, RA, and pericardium, which were approximately 0.84 and 0.95, respectively. The main reason was that according to our designed workflow, the performance of LA- and RA-EAT would depend on the accuracy of LA and pericardium segmentation, as well as RA and pericardium segmentation, respectively. We believed that the automatic segmentation of LA- and RA-EAT could assist future AF-related studies to avoid time-consuming and observer-dependent manual labeling.

## Limitations

This study had several limitations. First, the cardiac CT images were only collected from a single center, comprising two manufacturers’ CT scanners. This indicated that the degree of heterogeneous data required to be increased. We will use CT scans from multiple centers and different types of CT scanners to improve the adaptability of our deep learning models in the future. Second, the manual delineation of all cardiac structures was adopted by the consensus annotation from two experts. In the future, the inter-rater agreement will be used to validate the manual segmentation results. Third, the patients included in this study were those who suffered from AF, not normal volunteers or patients with other cardiovascular diseases. The contrast-enhanced cardiac CT images from patients with other heart diseases such as dilated cardiomyopathy or myxoma were obtained poor segmentation results by 3D U-Net models. After labeling the new data from other heart diseases, the transfer learning will be used to fine-tune our developed neural networks to improve the extensiveness of use.

## Conclusions

This study proposed a workflow for automatic segmentation of LA, RA, pericardium and EAT using deep learning on cardiac CT images in AF patients. Furthermore, we firstly developed the automatic segmentation of LA- and RA-EAT to prevent time-consuming manual delineation. In clinical practice, this workflow can assist in improving work efficiency and facilitating future applications in the prediction of AF recurrence.

## Data Availability

All data produced in the present study are available upon reasonable request to the authors.

## References

[1] R. S. Wijesurendra and B. Casadei, “Mechanisms of atrial fibrillation,” Heart, vol. 105, pp. 1860–1867, Dec 2019.

[2] M. Conte, L. Petraglia, S. Cabaro, V. Valerio, P. Poggio, E. Pilato, et al., “Epicardial Adipose Tissue and Cardiac Arrhythmias: Focus on Atrial Fibrillation,” Front Cardiovasc Med, vol. 9, p. 932262, 2022.

[3] A. Di Cori, G. Zucchelli, L. Faggioni, L. Segreti, R. De Lucia, V. Barletta, et al., “Role of pre-procedural CT imaging on catheter ablation in patients with atrial fibrillation: procedural outcomes and radiological exposure,” J Interv Card Electrophysiol, vol. 60, pp. 477–484, Apr 2021.

[4] S. Sarin, C. Wenger, A. Marwaha, A. Qureshi, B. D. Go, C. A. Woomert, et al., “Clinical significance of epicardial fat measured using cardiac multislice computed tomography,” Am J Cardiol, vol. 102, pp. 767–71, Sep 15 2008.

[5] S. H. Shin, M. Y. Park, W. J. Oh, S. J. Hong, H. N. Pak, W. H. Song, et al., “Left atrial volume is a predictor of atrial fibrillation recurrence after catheter ablation,” J Am Soc Echocardiogr, vol. 21, pp. 697–702, Jun 2008.

[6] F. M. Costa, A. M. Ferreira, S. Oliveira, P. G. Santos, A. Durazzo, P. Carmo, et al., “Left atrial volume is more important than the type of atrial fibrillation in predicting the longterm success of catheter ablation,” Int J Cardiol, vol. 184, pp. 56–61, Apr 1 2015.

[7] A. Njoku, M. Kannabhiran, R. Arora, P. Reddy, R. Gopinathannair, D. Lakkireddy, et al., “Left atrial volume predicts atrial fibrillation recurrence after radiofrequency ablation: a meta-analysis,” Europace, vol. 20, pp. 33–42, Jan 1 2018.

[8] Y. Akutsu, K. Kaneko, Y. Kodama, J. Suyama, H. L. Li, Y. Hamazaki, et al., “Association between left and right atrial remodeling with atrial fibrillation recurrence after pulmonary vein catheter ablation in patients with paroxysmal atrial fibrillation: a pilot study,” Circ Cardiovasc Imaging, vol. 4, pp. 524–31, Sep 2011.

[9] T. Takagi, K. Nakamura, M. Asami, Y. Toyoda, Y. Enomoto, M. Moroi, et al., “Impact of right atrial structural remodeling on recurrence after ablation for atrial fibrillation,” J Arrhythm, vol. 37, pp. 597–606, Jun 2021.

[10] J. Stojanovska, E. A. Kazerooni, M. Sinno, B. H. Gross, K. Watcharotone, S. Patel, et al., “Increased epicardial fat is independently associated with the presence and chronicity of atrial fibrillation and radiofrequency ablation outcome,” Eur Radiol, vol. 25, pp. 2298–309, Aug 2015.

[11] K. Nagashima, Y. Okumura, I. Watanabe, T. Nakai, K. Ohkubo, T. Kofune, et al., “Association between epicardial adipose tissue volumes on 3-dimensional reconstructed CT images and recurrence of atrial fibrillation after catheter ablation,” Circ J, vol. 75, pp. 2559–65, 2011.

[12] H. M. Tsao, W. C. Hu, M. H. Wu, C. T. Tai, Y. J. Lin, S. L. Chang, et al., “Quantitative analysis of quantity and distribution of epicardial adipose tissue surrounding the left atrium in patients with atrial fibrillation and effect of recurrence after ablation,” Am J Cardiol, vol. 107, pp. 1498–503, May 15 2011.

[13] Q. Tong, M. Ning, W. Si, X. Liao, and J. Qin, “3D Deeply-Supervised U-Net Based Whole Heart Segmentation,” ed, 2018, pp. 224–232.

[14] C. Payer, D. Štern, H. Bischof, and M. Urschler, “Multi-label Whole Heart Segmentation Using CNNs and Anatomical Label Configurations,” ed, 2018, pp. 190–198.

[15] L. Baskaran, G. Maliakal, S. J. Al’Aref, G. Singh, Z. Xu, K. Michalak, et al., “Identification and Quantification of Cardiovascular Structures From CCTA: An End-to-End, Rapid, Pixel-Wise, Deep-Learning Method,” JACC Cardiovasc Imaging, vol. 13, pp. 1163–1171, May 2020.

[16] S. Sharobeem, H. Le Breton, F. Lalys, M. Lederlin, C. Lagorce, M. Bedossa, et al., “Validation of a Whole Heart Segmentation from Computed Tomography Imaging Using a Deep-Learning Approach,” J Cardiovasc Transl Res, vol. 15, pp. 427–437, Apr 2022.

[17] H. H. Chen, C. M. Liu, S. L. Chang, P. Y. Chang, W. S. Chen, Y. M. Pan, et al., “Automated extraction of left atrial volumes from two-dimensional computer tomography images using a deep learning technique,” Int J Cardiol, vol. 316, pp. 272–278, Oct 1 2020.

[18] M. Abdulkareem, M. S. Brahier, F. Zou, A. Taylor, A. Thomaides, P. J. Bergquist, et al., “Generalizable Framework for Atrial Volume Estimation for Cardiac CT Images Using Deep Learning With Quality Control Assessment,” Front Cardiovasc Med, vol. 9, p. 822269, 2022.

[19] A. A. Mahabadi, B. Balcer, I. Dykun, M. Forsting, T. Schlosser, G. Heusch, et al., “Cardiac computed tomography-derived epicardial fat volume and attenuation independently distinguish patients with and without myocardial infarction,” PLoS One, vol. 12, p. e0183514, 2017.

[20] G. Milanese, M. Silva, L. Bruno, M. Goldoni, G. Benedetti, E. Rossi, et al., “Quantification of epicardial fat with cardiac CT angiography and association with cardiovascular risk factors in symptomatic patients: from the ALTER-BIO (Alternative Cardiovascular Bio-Imaging markers) registry,” Diagn Interv Radiol, vol. 25, pp. 35–41, Jan 2019.

[21] M. Masuda, H. Mizuno, Y. Enchi, H. Minamiguchi, S. Konishi, T. Ohtani, et al., “Abundant epicardial adipose tissue surrounding the left atrium predicts early rather than late recurrence of atrial fibrillation after catheter ablation,” J Interv Card Electrophysiol, vol. 44, pp. 31–7, Oct 2015.

[22] A. Hoori, T. Hu, J. Lee, S. Al-Kindi, S. Rajagopalan, and D. L. Wilson, “Deep learning segmentation and quantification method for assessing epicardial adipose tissue in CT calcium score scans,” Sci Rep, vol. 12, p. 2276, Feb 10 2022.

[23] T. Siriapisith, W. Kusakunniran, and P. Haddawy, “A 3D deep learning approach to epicardial fat segmentation in non-contrast and post-contrast cardiac CT images,” PeerJ Comput Sci, vol. 7, p. e806, 2021.

[24] C. F. Lu, F. T. Hsu, K. L. Hsieh, Y. J. Kao, S. J. Cheng, J. B. Hsu, et al., “Machine Learning-Based Radiomics for Molecular Subtyping of Gliomas,” Clin Cancer Res, vol. 24, pp. 4429–4436, Sep 15 2018.

[25] A. Bartoli, J. Fournel, L. Ait-Yahia, F. Cadour, F. Tradi, B. Ghattas, et al., “Automatic Deep-Learning Segmentation of Epicardial Adipose Tissue from Low-Dose Chest CT and Prognosis Impact on COVID-19,” Cells, vol. 11, Mar 18 2022.

[26] J. Nalepa, M. Marcinkiewicz, and M. Kawulok, “Data Augmentation for Brain-Tumor Segmentation: A Review,” Front Comput Neurosci, vol. 13, p. 83, 2019.

[27] H. S. Sacks and J. N. Fain, “Human epicardial adipose tissue: a review,” Am Heart J, vol. 153, pp. 907–17, Jun 2007.

[28] B. Franssens, H. Nathoe, T. Leiner, Y. Graaf, and F. Visseren, “Relation between cardiovascular disease risk factors and epicardial adipose tissue density on cardiac computed tomography in patients at high risk of cardiovascular events,” European Journal of Preventive Cardiology, vol. 24, 11/21 2016.

[29] Z. Liu, S. Wang, Y. Wang, N. Zhou, J. Shu, C. Stamm, et al., “Association of epicardial adipose tissue attenuation with coronary atherosclerosis in patients with a high risk of coronary artery disease,” Atherosclerosis, vol. 284, pp. 230–236, May 2019.

[30] T. D. Wang, W. J. Lee, F. Y. Shih, C. H. Huang, Y. C. Chang, W. J. Chen, et al., “Relations of epicardial adipose tissue measured by multidetector computed tomography to components of the metabolic syndrome are region-specific and independent of anthropometric indexes and intraabdominal visceral fat,” J Clin Endocrinol Metab, vol. 94, pp. 662–9, Feb 2009.

[31] A. A. Taha and A. Hanbury, “Metrics for evaluating 3D medical image segmentation: analysis, selection, and tool,” BMC Med Imaging, vol. 15, p. 29, Aug 12 2015.

[32] F. Commandeur, M. Goeller, A. Razipour, S. Cadet, M. M. Hell, J. Kwiecinski, et al., “Fully Automated CT Quantification of Epicardial Adipose Tissue by Deep Learning: A Multicenter Study,” Radiol Artif Intell, vol. 1, p. e190045, Nov 27 2019.

[33] X. He, B. J. Guo, Y. Lei, T. Wang, Y. Fu, W. J. Curran, et al., “Automatic segmentation and quantification of epicardial adipose tissue from coronary computed tomography angiography,” Phys Med Biol, vol. 65, p. 095012, May 11 2020.

