## Supplemental Materials for "Deep Learning-based Workflow for Automatic Extraction of Atria and Epicardial Adipose Tissue on cardiac Computed Tomography in Atrial Fibrillation"

**2D U-Net architecture and training parameters**

We developed a 2D U-Net model to perform the semi-automatic segmentation with 30 patients including 9600 images as training set and 8 patients including 2560 images as testing set. The cardiac CT and ROI images with a matrix size of 400 × 400 × 320 were fed into the 2D neural network and trained slice-by-slice.

The architecture of 2D U-Net model was similar to 3D model with few modifications (**Figure S1**). The 2D U-Net replaced all 3D kernels by 2D kernels such as 3 × 3 and 1 × 1 convolutional kernels. The kernel size of max-pooling operations and transposed convolutional kernels was converted to 5 × 5 and 3 × 3 with a stride of 2. The cross entropy was used as the loss function.

During the training progress, we used the stochastic gradient descent for updating weights with a momentum parameter of 0.9. Training parameters for LA and RA models were: learning rate = 0.01, mini-batch size = 2, max epoch = 8, learning rate drop period = 10 and learning rate drop factor = 0.3. It took about 140 minutes to train 2D U-Net models for LA and RA. The training parameters for pericardium segmentation using 2D U-Net were identical to those for LA and RA segmentation expect for the number of epochs. The max epoch for pericardial 2D segmentation was changed to 4 to prevent overfitting. It took about 45 minutes to train 2D segmentation model of pericardium.

**Performance of 2D U-Net models**

Dice coefficients assessed by 2D U-Nets for LA, RA and pericardium segmentation were 0.929 $\pm$ 0.019, 0.895 $\pm$ 0.054 and 0.944 $\pm$ 0.012, respectively. The 2D U-Nets for LA, RA and pericardium segmentation achieved the sensitivity of 0.921 $\pm$ 0.046, 0.858 $\pm$ 0.093 and 0.962 $\pm$ 0.010, and precision of 0.940 $\pm$ 0.039, 0.941 $\pm$ 0.013 and 0.927 $\pm$ 0.019. Most of the revision time spent on erasing the pericardial contours in pericardium segmentation, while filling in the missing regions in LA and RA segmentation. **Figure S2** shows the regions of 2D automatic segmentation that required to be correct.

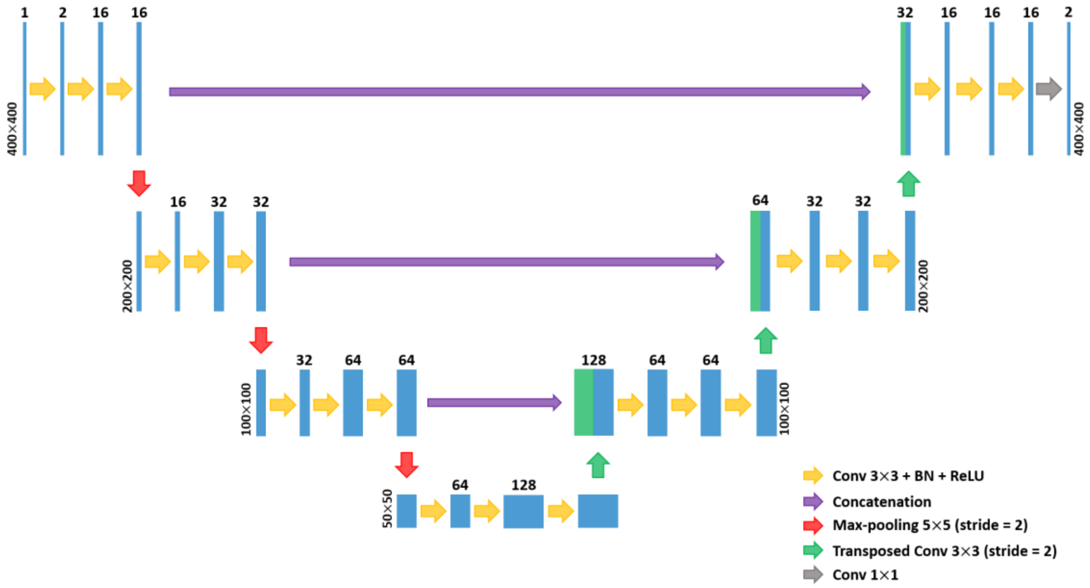


**Figure S1. 2D U-Net architecture.** The 2D U-Nets are used to assist in achieving a rapid delineation.

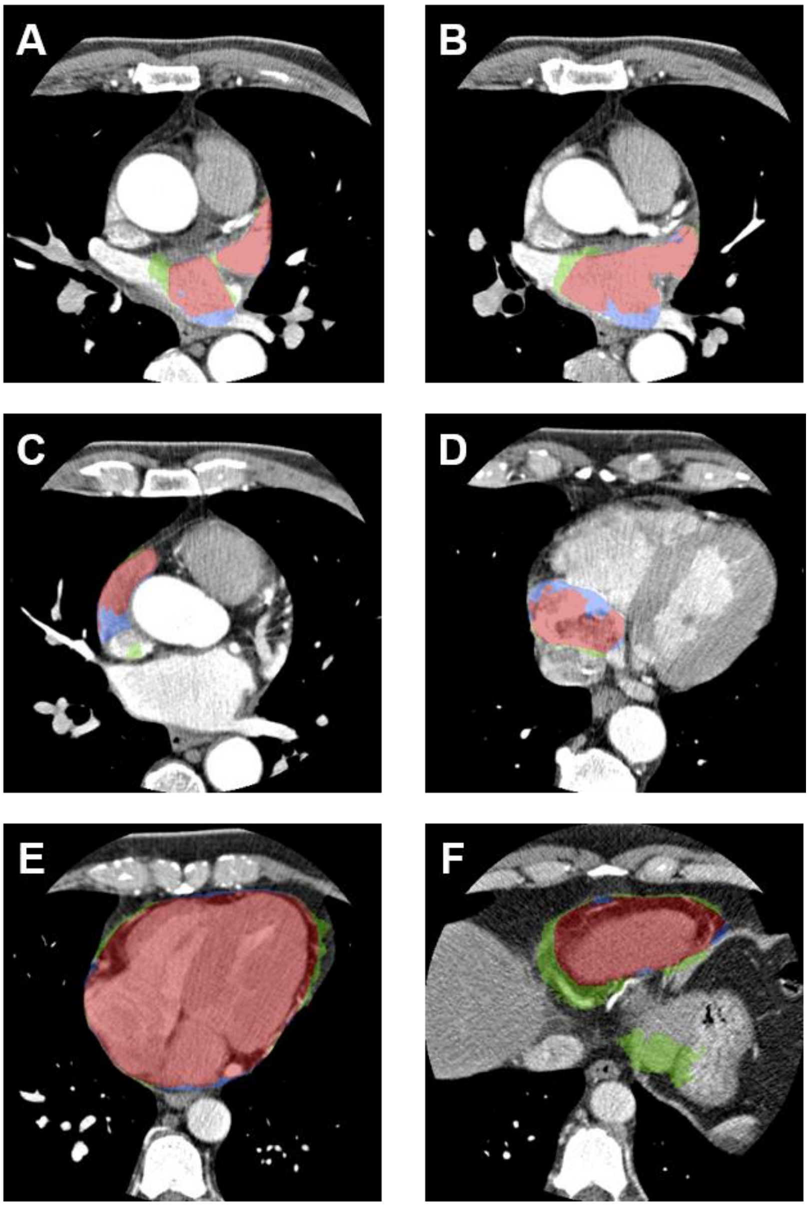


**Figure S2. Revised regions using 2D U-Net-assisted labeling.** LA ROIs are corrected the junctions of LA and PVs (A & B). RA ROIs are corrected the RA-SVC (C) and RA-IVC (D) junctions. Contours of pericardium ROIs are checked in each slice (E). Major revisions of pericardium ROIs are located around the inferior ventricle (F). Blue and green colors denote the manual delineation and 2D segmentation, respectively. Red color is the overlapping area of blue and green regions.

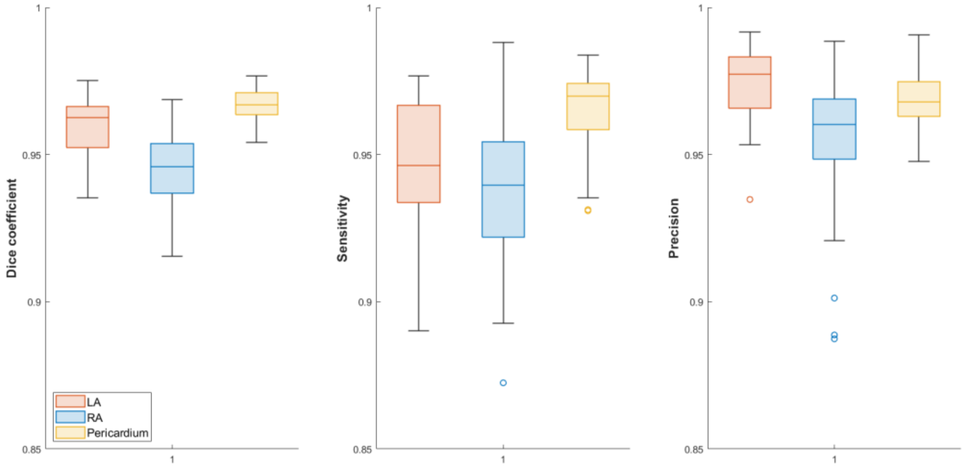


**Figure S3. Box plots of Dice coefficients, sensitivity and precision for LA, RA and pericardium assessed by 3D U-Net models.**
